# Pharmacovigilance of Integrase Strand Transfer Inhibitors in Botswana: Safety Signals, Report Completeness, and Causality Assessment

**DOI:** 10.64898/2026.09.08.26362506

**Authors:** Pono Pono, Vicky Cheng, Alan M. Jones

## Abstract

Integrase strand transfer inhibitors (INSTIs) are the preferred first-line anchor drug for antiretroviral therapy globally. However, post-marketing authorisation studies continue to find previously unidentified adverse events (AEs) associated with INSTIs. A pharmacovigilance study was conducted to investigate AEs associated with INSTIs submitted to the Botswana national pharmacovigilance database (BoMRA).

All suspected AE reports in which INSTIs were identified as the primary suspect were included for analysis. Descriptive analysis was used to characterise the AEs, and disproportionality analysis was used for signal detection. Causality assessment was conducted using the World Health Organization-Uppsala Monitoring Centre system, the Liverpool ADR Causality Assessment Tool, and the Naranjo Causality Assessment algorithm. Agreement between the methods was assessed through the Cohen’s Kappa (κ) test.

A total of *n* = 112 AE reports were reported for cabotegravir and dolutegravir as the ‘primary suspect’ drugs. Most reports involved females and adults, while dolutegravir was associated with the majority (78.6%). Skin and subcutaneous AEs (25.0%) and rash (10.7%) were the most frequently reported AEs at the system organ class (SOC) and reported preferred term (PT) levels, respectively. Reporter qualification influenced report completeness, with nurses submitting reports with the highest median completeness score.

Most reported AEs were consistent with the established safety profiles of cabotegravir and dolutegravir. However, the study identified unlabelled AEs for both cabotegravir and dolutegravir, including decreased libido, hallucination, severe obesity, acquired lipodystrophy, gynecomastia, cheilitis, and hyperpigmentation. These are potential safety signals that warrant further investigation and underscore the importance of national pharmacovigilance data for ongoing post-marketing safety surveillance and for generating local safety data insights.

## Introduction

Botswana is among the countries severely affected by human immunodeficiency virus (HIV), with a national prevalence rate of 20.8% [1]. Despite a high disease burden, it has achieved notable progress in HIV management. In 2025, Botswana was awarded Gold Tier certification for elimination of mother-to-child transmission (EMTCT) by the World Health Organization (WHO), becoming the first country globally to reduce mother-to-child HIV transmission to fewer than 250 per 100,000 live births and to achieve at least 95% coverage for antenatal care, HIV testing, and antiretroviral therapy (ART) enrolment [2].

Integrase strand transfer inhibitors (INSTIs) are the preferred anchor drug of ART globally due to their favourable efficacy, safety profiles, and high genetic barrier to HIV resistance [3, 4]. In 2016, Botswana was the first African country to introduce dolutegravir-based regimens as the preferred first-line for HIV treatment [5], and recently, cabotegravir long-acting for pre-exposure prophylaxis (PrEP) among vulnerable groups [6]. Although INSTIs exhibit a good safety profile, emerging evidence from post-marketing and cohort studies has revealed several adverse events (AEs), including neuropsychiatric disorders [7–9], weight gain, metabolic AEs [10], and congenital anomalies [11]. These AEs may negatively affect treatment adherence, quality of life (QoL), and overall clinical outcome of people living with HIV (PLWH).

Pharmacovigilance (PV) plays an essential role in the detection, assessment, understanding, and prevention of AEs and other medicine-related problems, thereby supporting regulatory decision-making and safeguarding public health [12]. Once medicinal products are approved, they undergo continuous post-marketing safety surveillance to ensure the risk-benefit ratio remain favourable throughout the product life cycle. Healthcare professionals (HCPs), patients, caregivers, and marketing authorisation holders (MAHs) contribute towards this initiative by reporting AEs to national PV centres [13]. The spontaneous reporting system (SRS) remains a cornerstone of post-marketing safety surveillance of medicines [14]. At the national level, PV centres maintain databases of spontaneous reports of suspected AEs, which contribute to the global database (VigiBase), maintained by the Uppsala Monitoring Centre, Program for International Drug Monitoring (PIDM) on behalf of WHO [15]. As the volume of reports to SRSs has continued to increase over the years, data-mining approaches such as disproportionality analysis are increasingly used to identify potential safety signals, which are later validated through controlled studies [4]. Despite progress in high-income countries, PV progress in low- and middle-income countries (LMICs) has lagged due to under-reporting, poor-quality spontaneous reports, insufficient stakeholder coordination, financial constraints, and limited skilled human resources [16, 17].

In Botswana, the Botswana Medicines Regulatory Authority (BoMRA) coordinates PV activities [18]. BoMRA established a decentralised national PV system in 2018 that is fully integrated into public health programmes (PHPs), including the national Human Immunodeficiency Virus (HIV) program and the Expanded Programme on Immunisation (EPI). Through collaboration with healthcare facilities, BoMRA receives and assesses AEs, records them in the national PV database (VigiFlow), and subsequently submits them to the global database, VigiBase [19]. However, analysis of national safety data remains limited. Therefore, the objectives of this retrospective analysis were to gain insights into the characteristics and patterns of AEs associated with INSTIs, to identify safety signals, to evaluate report completeness, and to conduct a causality assessment of the AE reports.

## Methods

### Study design

A retrospective, descriptive, cross-sectional study was conducted using spontaneous AE reports extracted from the Botswana national PV database. The analysis included spontaneously reported AEs associated with INSTIs reported as the primary suspect drug.

### Data extraction

All spontaneous AE reports reported from 1^st^ January 2013 to 3^rd^ July 2026 were extracted from the Botswana national PV database on the 3^rd^ of July 2026. A comma-separated values (CSV) file was downloaded and processed in RStudio version 4.5.0 (2024.12.0 Build 467). The CSV file contains four main sheets: i) Cases, ii) Drugs, iii) Reactions, and iv) Drug-Reaction link. The Cases sheet contains the following variables: Safety report ID, completeness score, reporter qualification, sex, age, age group, seriousness, and seriousness criteria. Drugs: Safety report ID, WHODrug active ingredient variant, role, indication, start date, end date of treatment, and action taken with the drug. Reactions: Safety report ID, Mapped term, MedDRA preferred term, start date, end date of reaction, and outcome. Drug-reaction link: Safety report ID, Time-to-onset (TTO), whether dechallenge was performed, reaction resolved or resolving, rechallenge performed, reaction recurred or not. The four data files were combined to generate a Masterfile; safety report ID was used as an anchor to horizontally merge the sheets. Furthermore, the safety report ID was used to deduplicate the Masterfile.

### Descriptive analysis

Only AE reports in which an INSTI was identified as the “primary suspect” drug were included in the analysis. For this analysis, cabotegravir and cabotegravir sodium were aggregated into a single cabotegravir category, and dolutegravir and dolutegravir sodium were combined into a single dolutegravir category because they are different salt forms of the same active pharmaceutical ingredient (API). Descriptive statistics were used to summarise the demographic and clinical profiles of ICSRs, and these are presented as frequencies (*n*) and proportions (*%*).

### Signal detection

Disproportionality analysis to detect safety signals associated with INSTIs at the preferred-term level was conducted, using the entire database as the comparator. The signals of disproportionate reporting (SDRs) were generated by employing the Bayesian confidence propagation neural network (BCPNN). BCPNN was selected because it provides a Bayesian shrinkage estimate that stabilises the disproportionality measure. Furthermore, it is robust in cases of sparse and small-sample databases [20, 21]. The BoMRA database contains 9,647 AE reports. Therefore, other disproportionality analysis methods, such as reporting odds ratios (ROR) and proportional reporting ratios (PRR), would produce inflated or unstable estimates, thus providing false positives. BCPNN is based on a two-by-two contingency table (Supporting information Table S1). A positive safety signal was identified when the number of observed PTs was ≥ 3 and IC_025_ > 0 (Supporting information Table S2).

### Causality assessment: case-by-case analysis

Causality assessment was conducted to evaluate the likelihood that the drug of interest led to the reported suspected AEs. All AE reports were downloaded as PDF files, and the data were extracted into tabular format in Excel. The extracted data included administrative information, patient’s demographics, drugs (suspected and concomitant), seriousness and fatal information, reactions/events (including any laboratory investigations), a narrative case summary, and further information. Medication errors and AE reports with insufficient information were excluded.

The World Health Organization-Uppsala Monitoring Centre (WHO-UMC) system [22], the system, the Liverpool ADR Causality Assessment Tool (LCAT) [23, 24] and the Naranjo Causality Assessment algorithm [25] were used in a case-by-case analysis to establish a link between each adverse event and the suspected drug. Furthermore, agreement between the methods was assessed using Cohen’s kappa coefficient.

### Statistics

Continuous variables were summarised using medians and interquartile ranges, while categorical variables were described using frequencies and percentages. Differences in completeness scores between reporter groups were assessed using the Kruskal-Wallis test, followed by pairwise Wilcoxon rank-sum tests with Benjamini-Hochberg adjustment [26]. Logistic regression was used to identify predictors of seriousness and completeness of AE reports. Statistical significance was defined as *p* < 0.05. All analyses were performed using RStudio version 4.5.0 (2024.12.0 Build 467), © 2009-2024 Posit Software, PBC.

## Results

### Clinical and demographic characteristics of the adverse events

A total of *n* = 9,647 AE reports were retrieved from the database. Among these, *n* = 112 AE reports were reported when one of the INSTIs was listed as a ‘primary suspect’ (Figure *1*). Table 1 shows clinical and demographic characteristics of AEs associated with INSTIs. Among the 112 identified AE reports, the majority were reported in females (*n* = 69; 61.6 %). The 18-44 years age group had the highest number of reports (*n* = 47; 42.0%), followed by the 45-64 years age group (*n* = 36; 32.1%), and the 28-day to 23-month age group had the lowest number of reports (*n* = 1; 0.9 %). Combined reports of dolutegravir and dolutegravir sodium accounted for the highest share (*n* = 87; 78.6%), followed by cabotegravir and cabotegravir sodium (*n* = 25; 11.7%). Among the AE reports, 76.8% were classified as non-serious, and 22.3% considered serious. For those AE reports that were classified as serious, 10.7% were for other medically important conditions, 3.6% were disabling/incapacitating and life-threatening, and 0.9% were fatal. Other healthcare professionals (nurses) submitted most AE reports (50.9%), followed by pharmacists (28.6%) and physicians (17.9%).

**Figure 1.**
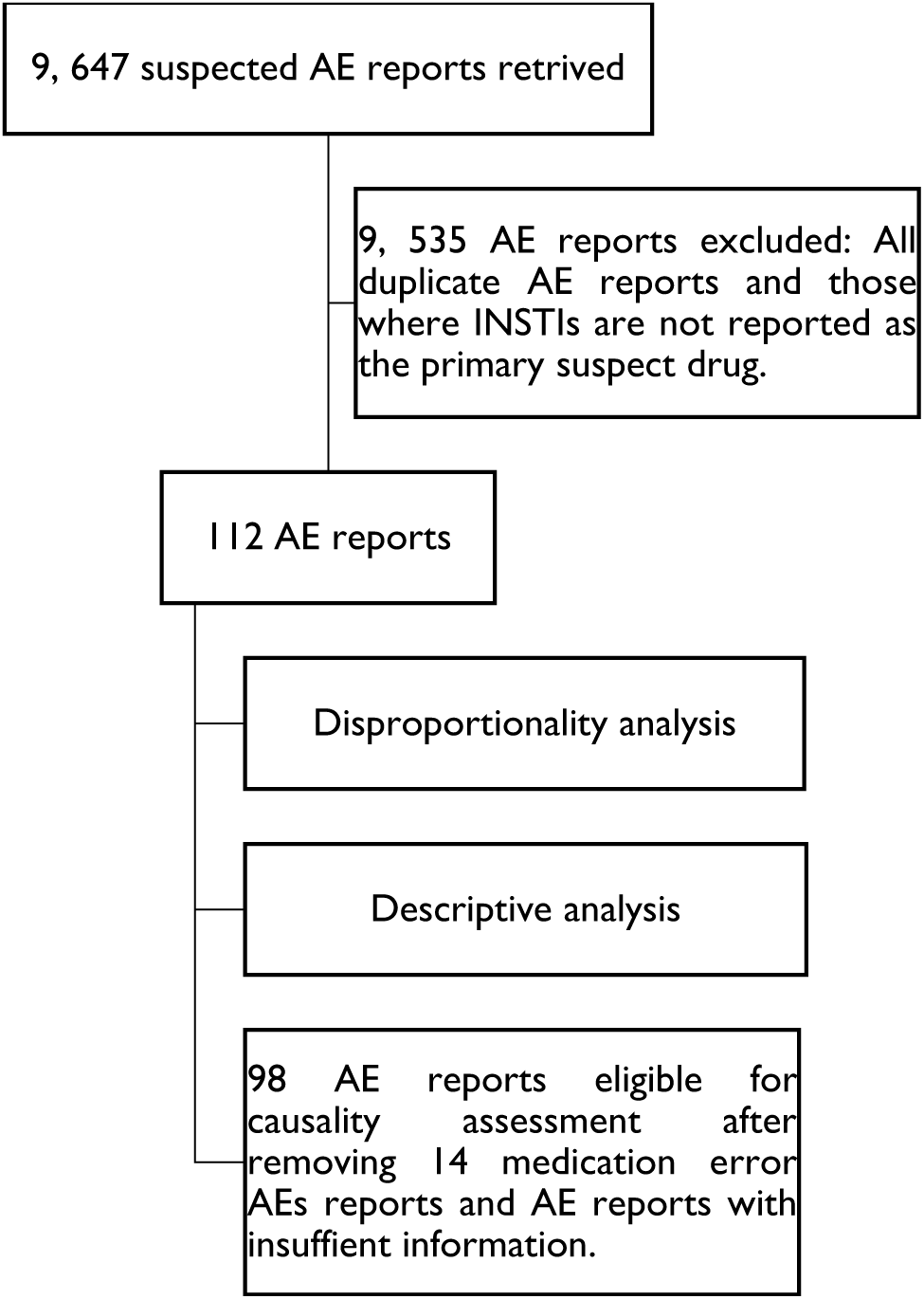
Data screening of all adverse events retrieved from the Botswana Medicines Regulatory Authority’s database.

**Table 1.** Clinical characteristics of patients who experienced adverse events associated with integrase strand transfer inhibitors.

| Variable | *n* | Percentage |
| --- | --- | --- |
| Patient sex |  |  |
| Female | 69 | 61.6% |
| Male | 38 | 33.9% |
| Unknown | 5 | 4.5% |
| Patient age |  |  |
| 28 days to 23 months | 1 | 0.9% |
| 2 - 11 years | 5 | 4.5% |
| 12 - 17 years | 1 | 0.9% |
| 18 - 44 years | 47 | 42.0% |
| 45 - 64 years | 36 | 32.1% |
| 65 - 74 years | 7 | 6.3% |
| ≥ 75 years | 4 | 3.6% |
| Unknown | 11 | 9.8% |
| Drug |  |  |
| Dolutegravir | 81 | 72.3% |
| Cabotegravir | 17 | 15.2% |
| Cabotegravir sodium | 8 | 7.1% |
| Dolutegravir sodium | 6 | 5.4% |
| Serious |  |  |
| Yes | 25 | 22.3% |
| No | 86 | 76.8% |
| Unknown | 1 | 0.9% |
| Seriousness criteria |  |  |
| Death | 1 | 0.9% |
| Life threatening | 4 | 3.6% |
| Caused/prolonged hospitalization | 3 | 2.7% |
| Disabling/incapacitating | 4 | 3.6% |
| Congenital anomaly/birth defect | 3 | 2.7% |
| Other medically important conditions | 12 | 10.7% |

| Reporter qualification |  |  |
| --- | --- | --- |
| Physician | 20 | 17.9% |
| Pharmacist | 32 | 28.6% |
| Other Health Professionals (nurses) | 57 | 50.9% |
| Consumer/Non-Health Professional | 1 | 0.9% |
| Unknown | 2 | 1.8% |

### Descriptive analysis of suspected adverse events associated with integrase strand transfer inhibitors

The most common system organ class (SOC) involved in AEs was skin and subcutaneous tissue disorders (25.0%), followed by renal and urinary disorders (18.8%) and general disorders and administration site conditions (17.9%), whereas ear and labyrinth disorders were the least frequently reported SOC (0.9%) (Figure *2*). At the preferred term level, the top 6 reported preferred terms were rash (10.7%), renal failure (9.8%), pain (8.0%), pruritus (8.0%), renal impairment (7.1%), and injection site pain (6.3%) (Table *2*).

**Figure 2.**
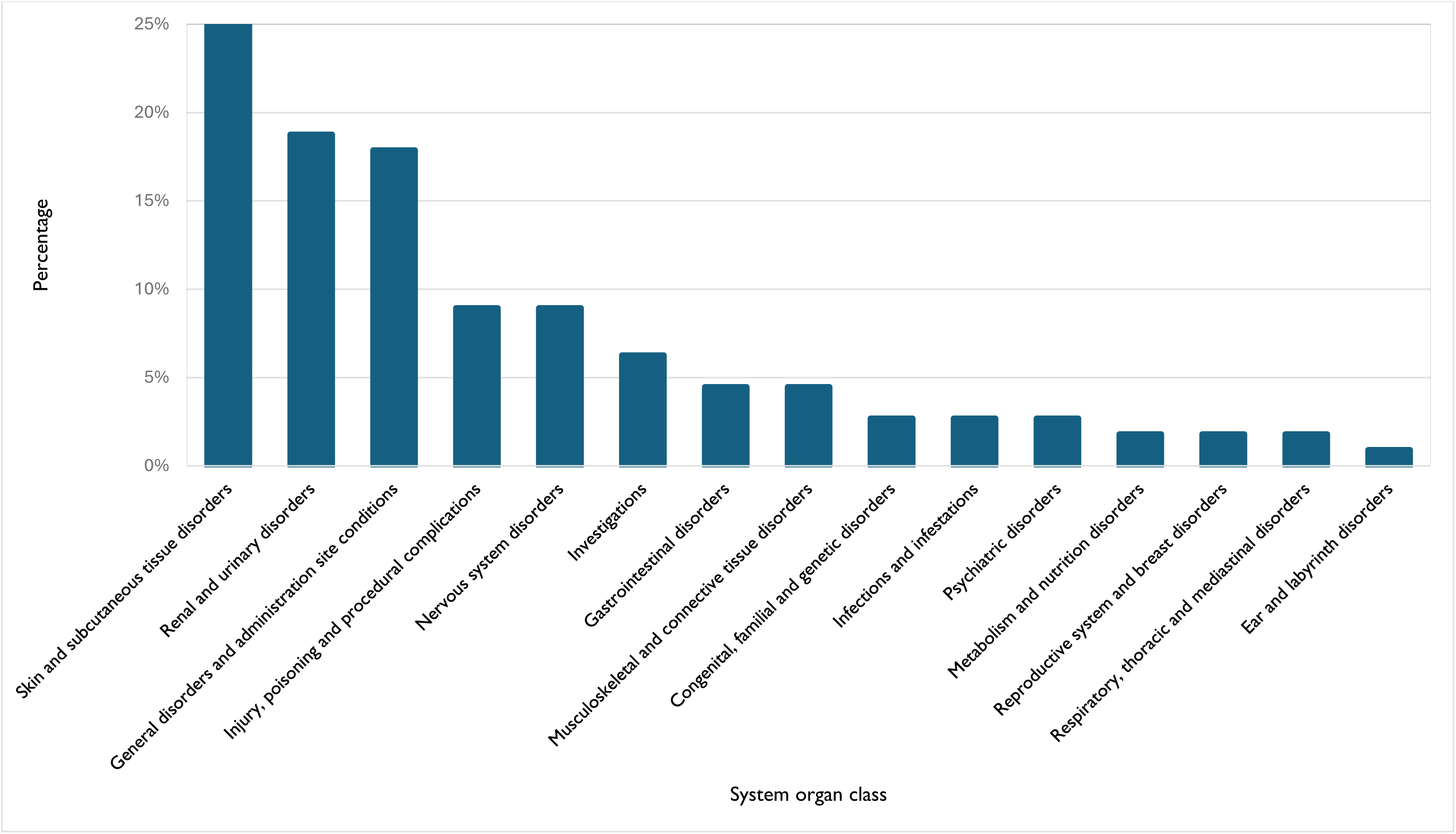
Distribution of suspected adverse events (AEs) associated with INSTIs at the system organ class (SOC) level. A total of 112 AE reports were included in the analysis; the percentages represent the proportion of each SOC.

**Table 2.** Classification of suspected adverse event (AEs) associated with INSTIs preferred term (PT) level. A total of 112 AE reports were included in the analysis. The percentage represents the proportion of each PT.

| Preferred term | Count | Percentage |
| --- | --- | --- |
| Rash | 12 | 10.7% |
| Renal failure | 11 | 9.8% |
| Pain | 9 | 8.0% |
| Pruritus | 9 | 8.0% |
| Renal impairment | 8 | 7.1% |
| Injection site pain | 7 | 6.3% |
| Headache | 4 | 3.6% |
| Swelling | 4 | 3.6% |
| Lipodystrophy acquired | 4 | 3.6% |
| Creatinine renal clearance decreased | 3 | 2.7% |
| Dizziness | 3 | 2.7% |
| Medication error | 3 | 2.7% |
| Overdose | 3 | 2.7% |
| Pain in extremity | 3 | 2.7% |
| Neuropathy peripheral | 2 | 1.8% |
| Rash maculo-papular | 2 | 1.8% |
| Weight increased | 2 | 1.8% |
| Neural tube defect | 2 | 1.8% |
| Injection site swelling | 2 | 1.8% |

### Logistic regression

Logistic regression was used to assess potential predictors of the seriousness of AE reports. After adjustment for patient sex, age group, and INSTI drug, males were associated with significantly lower odds of serious AEs (*p* = 0.038). Age group was not significantly associated with seriousness (*p* = 0.353), and dolutegravir did not differ from cabotegravir in the odds of serious AEs (*p* = 0.845) (Table *3*).

**Table 3.** Multivariable logistic regression analysis of factors that affect the seriousness of the suspected AEs. Sex, age, and drug were included in the model as predictors of the seriousness of AE reports.

| Predictor | Category | Adjusted OR | 95% CI | p-value |
| --- | --- | --- | --- | --- |
| Sex | Male vs Female | 0.28 | 0.07 - 0.86 | 0.038* |
| Age group | Non-Adult vs Adult | 0.35 | 0.02 - 2.36 | 0.353 |
| Drug | Dolutegravir vs Cabotegravir | 1.13 | 0.35 - 3.89 | 0.845 |
| Notes: OR: Odds ratios; CI: Confidence interval. The outcome variable was the seriousness of the adverse event report (serious vs non-serious). Reference categories: female sex, adult age group, and cabotegravir. *$p < 0.05$. | | | | |

### Signal detection

Disproportionality analysis revealed 7 positive safety signals. The strongest signal was for cabotegravir, injection-site pain (*n* = 7, IC = 3.0, IC_025_;1.8), followed by swelling (*n* = 4, IC = 2.8, IC_025_; 1.0) and pain (*n* = 8, IC = 1.9, IC_025_; 0.7). For dolutegravir the strongest signals were overdose, (*n* = 3, IC= 2.6, IC_025_; 0.6) and rash (*n* = 8, IC= 1.3, IC_025_; 0.2) (Table *4*).

**Table 4.** Disproportionality analysis of adverse events associated with cabotegravir and dolutegravir at the preferred term level.

| Drug | PT | $n$ | $IC_{025}$ | IC |
| --- | --- | --- | --- | --- |
| Cabotegravir | Injection site pain | 7 | 1.8 | 3.0 |
| Cabotegravir | Swelling | 4 | 1.0 | 2.8 |
| Cabotegravir | Pain | 8 | 0.7 | 1.9 |
| Dolutegravir | Overdose | 3 | 0.6 | 2.6 |
| Dolutegravir | Rash | 8 | 0.2 | 1.3 |
| Dolutegravir | Medication error | 3 | 0.2 | 2.2 |
| Cabotegravir | Rash | 4 | 0.1 | 1.8 |
| Notes: PT; preferred term, IC; information component; $IC_{025}$; lower limit of 95% CI of the IC. Positive signals were detected when $IC_{025} > 0$ and $a \geq 3$. | | | | |

### Completeness of the submitted adverse event reports

The estimated median score for nurses was 0.70, compared with 0.47 for pharmacists and 0.56 for physicians (Supporting information Table S4). A comparison of reporter qualifications and report completeness (excluding consumers/non-healthcare professionals) revealed significant differences in scores. Relative to nurses, pharmacists and physicians had significantly lower completeness scores (adjusted *p* < 0.001) and (adjusted *p* = 0.046), respectively. However, there was no significant difference between AE reports submitted by pharmacists and physicians (adjusted *p* = 0.241) (Supporting information Table S5).

Linear regression of completeness score versus reporter qualification revealed that pharmacists and physicians, compared with nurses, had significantly lower scores (*p* < 0.001) and (*p* = 0.02), respectively (Supporting information Table S6). The seriousness of the AE reports was a predictor of completeness. Serious AE reports had a median completeness score of 0.72 (IQR: 0.57-1.00), compared with non-serious reports at 0.60 (IQR: 0.37-0.70) (Supporting information Table S7). Logistic regression demonstrated that higher completeness was associated with higher odds of the AE reports being classified as serious (OR = 11.38, 95% CI 1.75-84.97, *p* = 0.013) (Supporting information Table S8).

### Causality assessment

After removal of 14 AE reports, (*n* = 98) were eligible for causality assessment to determine the likelihood of a causal relationship between the reported AEs and the drugs (Figure 1). Across all three causality assessment methods, the majority of AEs reported for cabotegravir were classified as probable, with only one classified as definite. In contrast, most of the AEs associated with dolutegravir were classified as possible across all three causality assessment tools (Table 5).

**Table 5.** Causality assessment classification of 98 adverse events associated with cabotegravir and dolutegravir. Values are presented as frequencies (*n*) and percentages within each drug.

| Drug | Category | n | % |
| --- | --- | --- | --- |
| Liverpool ADR Causality Assessment Tool |  |  |  |
| Cabotegravir | Definite | 1 | 2% |
| Cabotegravir | Possible | 13 | 31% |
| Cabotegravir | Probable | 27 | 64% |
| Cabotegravir | Unlikely | 1 | 2% |
| Dolutegravir | Possible | 47 | 53% |
| Dolutegravir | Probable | 24 | 27% |
| Dolutegravir | Unlikely | 18 | 20% |
| The WHO-UMC system |  |  |  |
| Cabotegravir | Certain | 1 | 2% |
| Cabotegravir | Possible | 14 | 33% |
| Cabotegravir | Probable | 26 | 62% |
| Cabotegravir | Unlikely | 1 | 2% |
| Dolutegravir | Possible | 47 | 53% |
| Dolutegravir | Probable | 24 | 27% |
| Dolutegravir | Unassessable | 2 | 2% |
| Dolutegravir | Unlikely | 16 | 18% |
| The Naranjo Causality Assessment Algorithm |  |  |  |
| Cabotegravir | Definite | 1 | 2% |
| Cabotegravir | Possible | 9 | 21% |
| Cabotegravir | Probable | 32 | 76% |
| Dolutegravir | Possible | 65 | 73% |
| Dolutegravir | Probable | 24 | 27% |
| Notes: LCAT: Liverpool ADR Causality Assessment Tool; WHO-UMC: World Health Organization-Uppsala Monitoring Centre. |  |  |  |

The highest agreement was observed between the LCAT and the WHO-UMC system (*κ* = 0.951). In contrast, there was noticeable disagreement between the Naranjo algorithm and both the LCAT (*κ* = 0.655) and the WHO-UMC systems (*κ* = 0.628) (Supporting information Table S9).

### Unlabelled adverse events associated with cabotegravir and dolutegravir

A total of *n* = 19 AEs were not listed in the relevant summary of product characteristics (SmPCs) of cabotegravir and dolutegravir. Among these, 17 were associated with dolutegravir, and 2 were associated with cabotegravir. WHO-UMC system and the LCAT classified 12 AEs as possible, 5 AEs as unlikely, and 2 AEs as unassessable/unclassifiable. Both unassessable/ unclassifiable AE reports involved neural tube defects, for which there was no exposure date during pregnancy. Therefore, it was not possible to establish whether dolutegravir exposure occurred during the periconceptional period or early first trimester, as these are critical time periods for neural tube development (Table 6).

**Table 6.**
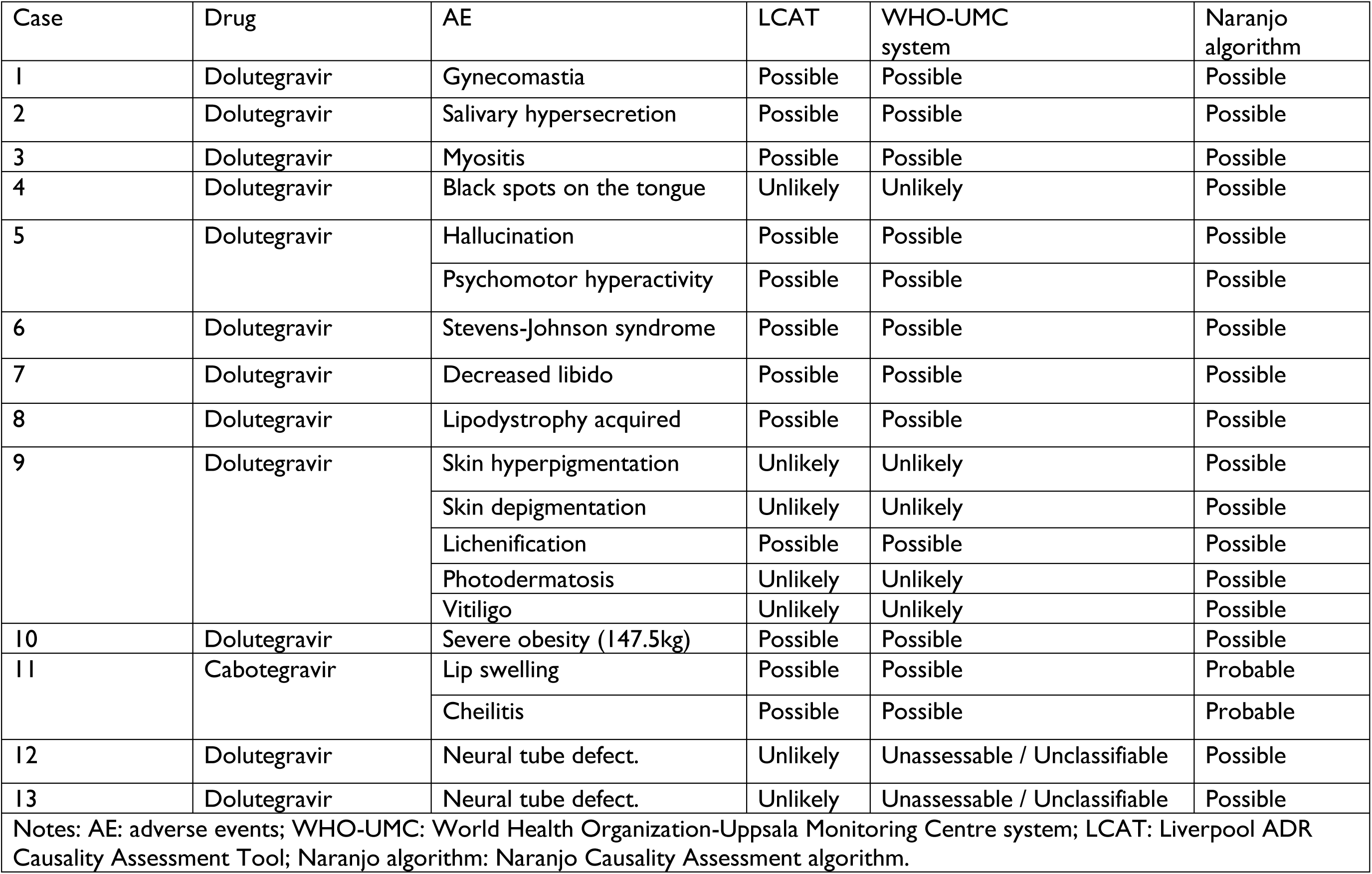
Adverse events not listed in the summaries of product characteristics of cabotegravir and dolutegravir.

## Discussion

This retrospective study reviews AEs associated with INSTIs in Botswana’s national pharmacovigilance database. Dolutegravir accounted for most AE reports. Cutaneous and renal AEs were most frequently reported at the SOC level, whereas rash, renal failure, and pruritus were most frequently reported at the PT level. Several AEs not listed on the dolutegravir SmPC were identified, including decreased libido, psychomotor hyperactivity and hallucination, severe obesity, acquired lipodystrophy, and gynecomastia. Disproportionality analysis revealed several potential safety signals for cabotegravir, including injection-site pain, swelling, and rash, whereas rash was associated with dolutegravir. Reporter qualification was a predictor of report completeness, and completeness was associated with serious AEs reports.

The predominance of AEs reports among females and individuals aged 18-44 years is consistent with HIV epidemiology in Botswana, where HIV prevalence is higher amongst adults (20.8%) and females (66.8%) [1]. Furthermore, dolutegravir accounted for most of the AE reports. In Botswana, dolutegravir-based regimen has been used as first line for HIV treatment since 2016 [5]. Therefore, a high number of dolutegravir-associated AEs does not necessarily reflect an unfavourable safety profile of dolutegravir but rather prescribing patterns and drug utilisation. Most of the AEs reported were classified as non-serious. However, female sex was significantly associated with higher odds of serious AEs compared to male sex. This aligns with evidence showing that females experience a higher frequency of AEs overall compared to males, potentially due to several factors, including differences in healthcare services utilisation, and sex-related differences in the pharmacokinetic and pharmacodynamic effects of the drug [27].

Most of the AE reports were submitted by HCPs, demonstrating active participation in medicines safety monitoring and reflecting successful integration of pharmacovigilance into routine clinical practice. Among HCPs, nurses submitted the most AEs, followed by physicians and pharmacists. Botswana has a decentralised HIV management model where nurse prescribers provide the majority of HIV care, particularly in healthcare facilities with limited physician availability [2]. Furthermore, nurses represent a larger proportion of all HCPs in Botswana, which may explain reporting patterns in the current study [2].

Reporter qualification was significantly associated with report completeness, with nurses submitting reports that had a higher median completeness score than those submitted by other reporters. A significant difference was noted between reports submitted by nurses and those submitted by pharmacists and physicians. However, no significant differences were noted between pharmacists and physicians. This may be attributed to the central role nurses play in delivering HIV care services in Botswana [28]. Similar observations were reported in pharmacovigilance studies in Germany and South Africa, where report quality varied across the reporters [29, 30]. These findings suggest that professional background, clinical responsibility, and familiarity with pharmacovigilance reporting pathways may influence the quality and frequency of AE reports submitted.

Report completeness was significantly associated with the seriousness of the AE, with serious AE reports exhibiting higher completeness. These findings are consistent with previous studies from SRS databases in the United States, the United Kingdom, Germany, France, Denmark, and South Africa [29–31]. Serious AEs are most likely to receive immediate attention, a detailed investigation, and closer patient monitoring, resulting in detailed information [32, 33]. Although definitive causality cannot be established from spontaneous reports only, AE reports with higher completeness scores may enhance the robustness and reliability of causality assessment.

The disproportionality analysis revealed four safety signals associated with cabotegravir, including injection site pain, swelling, rash, and pain. Among these, injection site reactions (ISRs) were most frequently reported. Cabotegravir long-acting is formulated as a nanosuspension for *intra*muscular injection, which may cause local inflammation, potentially explaining the higher frequency of ISR AEs. Clinical trials and post-marketing surveillance studies have consistently reported injection site pain as the most reported PT [34, 35].

Cutaneous AEs were the most frequently reported reactions in the current study, with pruritus and rash reported for both cabotegravir and dolutegravir. Furthermore, disproportional analysis also identified rash as a safety signal for both cabotegravir and dolutegravir. Cutaneous reactions are recognised AEs for both cabotegravir and dolutegravir [36, 37]. These AEs are thought to be immune-mediated hypersensitivity reactions and are usually mild to moderate in severity, typically presenting as generalised maculopapular eruptions [38]. However, the current study identified a case of Stevens-Johnson syndrome associated with dolutegravir and was classified as possible across three causality assessment methods. Similarly, dolutegravir-induced Stevens-Johnson syndrome has been reported in Sri Lanka [39]. In addition, other cutaneous and hypersensitivity-related AEs not listed in the relevant SmPCs were identified, including lip swelling, cheilitis, skin pigmentation, skin hyperpigmentation, vitiligo, lichenification, and photodermatosis. Clinical trials have reported rashes and hypersensitivity reactions among individuals receiving a dolutegravir-based regimen [40]. The occurrence of cutaneous AEs for both cabotegravir and dolutegravir may suggest an INSTI class effect.

Renal and urinary AEs were the second most reported at the SOC level, with several PTs, including reduced creatinine renal clearance, renal failure, and renal impairment. Although most reported AE reports were associated with dolutegravir, this should be interpreted with caution, as spontaneous reports alone cannot confirm causality. Dolutegravir is known to contribute to a benign increase in serum creatinine and decrease creatinine clearance during the first to fourth week after treatment initiation [41]. Therefore, this does not reflect the nephrotoxicity of dolutegravir but its inhibitory effects on the multidrug and toxin extrusion transporter 1 (MATE1), organic cation transporter 2 (OCT2), and multidrug and toxin extrusion transporter 2-K (MATE2-K) [42]. However, the observed renal AEs may partly reflect the contribution of concomitant ART, particularly tenofovir disoproxil fumarate, which is a component of the HIV first-line combination in Botswana. Tenofovir disoproxil fumarate has a well-established association with renal toxicity as well as reduced renal function [43]. Therefore, there is a need for continued renal monitoring among patients receiving tenofovir disoproxil fumarate and dolutegravir-containing regimens within routine clinical practice.

The current study identified neuropsychiatric AEs not listed in the dolutegravir SmPC, including decreased libido, psychomotor hyperactivity, and hallucination. Dolutegravir has been associated with several psychiatric AEs in clinical trials and observational studies, including insomnia, abnormal dreams, anxiety, and depression [44]. Dolutegravir crosses the blood-brain barrier (BBB) and reaches measurable concentrations exceeding the *in vitro* IC_50_ and the therapeutic concentration of 0.2 ng/mL and 2.4 ng/mL, respectively [45]. Montenegro-Burke et al. demonstrated that dolutegravir disrupts cerebral metabolic pathways, leading to reactive stress and mitochondrial dysfunction, which may be a plausible mechanistic explanation for clinical psychiatric manifestations associated with dolutegravir [46]. The current study classifies both psychomotor hyperactivity and hallucination as possible across all three causality assessment methods, suggesting a possible causal relationship. Furthermore, decreased libido was classified as possible in the current study. Observational studies have previously reported erectile dysfunction and transient loss of sexual desire in males receiving a dolutegravir-based regimen [47, 48]. Therefore, continued psychiatric AEs monitoring is warranted for patients on dolutegravir-based regimens as they may impede treatment adherence.

Metabolic and adipose tissue AEs identified in the current study included severe obesity, acquired lipodystrophy, and gynecomastia, all of which were classified as possible by the causality assessment methods. This is consistent with growing evidence suggesting an association between dolutegravir and weight gain and alterations in fat body composition. Observational studies have reported weight gain among individuals receiving a dolutegravir-based regimen [49–51], with females and people of African and Hispanic ancestry emerging as a high-risk population [52]. Several biological mechanisms have been proposed, including disruption of hunger signalling, leptin suppression, adipocyte fibrosis, inhibition of human melanocortin 4 receptors, and mitochondrial dysfunction [53]. Therefore, severe obesity and lipodystrophy identified in the study may be biologically plausible. However, severe obesity and lipodystrophy are multifaceted and may be influenced by several risk factors, including the “return to health” phenomenon after initiation of ART, concomitant ARTs, HIV-immunologic and microbiome-mediated weight changes, age, and diet [53]. Nevertheless, these findings reinforce the continued need to monitor weight and metabolic parameters in individuals on a dolutegravir-based regimen.

Causality assessment classification revealed a consistent pattern across all three causality assessment methods. Most AEs associated with cabotegravir were classified as ‘Probable’, most likely because the majority of the AEs were ISRs and exhibited a strong temporal relationship with the administration of the long-acting cabotegravir. In contrast, most AEs associated with dolutegravir were classified as ‘Possible.’ Many of these AE reports contained potential confounding factors and insufficient clinical information to support a stronger causal classification, resulting in most being classified as ‘Possible.’

There was a high level of agreement between the WHO-UMC system and LCAT, indicating similar classification of the AEs. In contrast, there was lower agreement between the Naranjo algorithm and other causality assessment methods. The differences between the Naranjo algorithm and other causality assessment methods could be attributed to methodological differences. The Naranjo algorithm did not classify AEs as ‘doubtful’ because all AEs were allocated 2 points for the second question, since all suspected AEs occurred after drug administration. While the LCAT and the WHO-UMC system classified some AE as ‘Unlikely.’

Previous studies have reported similar outcomes. Mittal et al. [54] and Acharya et al. [55] found moderate agreement between the WHO-UMC system and the Naranjo algorithm (κ = 0.70) and (κ = 0.60), respectively, which corroborates aspects of the current study. However, Gupta et al. [56] reported diverging outcomes from the current study. Gupta and colleagues reported that the three methods commonly classify most AEs as ‘Probable’. Furthermore, negative and poor agreement (κ = −0.16) between the WHO-UMC system and LCAT was reported [56].

Several limitations of the study are acknowledged. First, only reports in which INSTIs were reported as the primary suspect drug were included in the analysis, resulting in a small sample size of 112 AE reports. This limited the ability to detect rare AEs, conduct subgroup analyses, and investigate AE reports in which INSTIs are concomitant medications. Second, only 25 serious AE reports were included in the regression analysis, which may have reduced statistical precision and resulted in wider confidence intervals. Third, several AE reports lacked clinical information, particularly medical history, treatment dates, and concomitant medications, which hindered a detailed assessment of causality. Fourth, underreporting, reporting bias, and variable report quality are inherent limitations of SRSs that may influence the frequency and patterns of reported AEs. Furthermore, SRSs lack a denominator, preventing calculation of incidence rates and estimation of relative risks. Finally, ART is administered as a combination; therefore, patients receiving dolutegravir were also exposed to concomitant medications, such as tenofovir disoproxil fumarate. Hence, the influence of concomitant medications and other confounding factors cannot be excluded.

This study identified several AEs not listed in the relevant SmPCs, highlighting the complementary role of pharmacovigilance in detecting rare, delayed-onset, and population-specific AEs that may not be identified during pre-market authorisation clinical trials. These exploratory findings do not establish causality. However, they provide important real-world safety evidence that will inform future cohort, pharmacoepidemiological, and mechanistic studies. Furthermore, this work demonstrates the importance of Botswana’s national PV systems in generating local safety data that may inform future medicine safety surveillance, risk-management strategies, and regulatory decision-making.

## Supplementary Information

Table S1. Two-by-two contingency table. Table S2. Algorithms used for signal detection.

Table S3. Disproportionality analysis of cabotegravir and dolutegravir at the preferred term level.

Table S4. Completeness score by reporter qualification

Table S5. Pairwise Wilcoxon Comparisons with Benjamini-Hochberg (BH) adjustment of completeness scores by reporter

Table S6. Linear regression of completeness score versus reporter qualification Table S7. Linear regression of completeness score versus seriousness

Table S8. Logistic regression of completeness score versus seriousness

Table S9. Agreement statistics between the WHO-UMC system, the Liverpool ADR Causality Assessment Tool (LCAT), and the Naranjo Causality Assessment algorithm.

## Acknowledgements

The authors thank the Botswana Medicines Regulatory Authority for providing the data for this study.

## Author contribution

PP: Conceptualisation, Methodology, Data curation, and Writing-original draft. AJ & VC: Methodology, Conceptualisation, Validation, Supervision, and Writing – review & editing. All authors contributed to and approved the manuscript.

## Funding

This research was conducted as part of a PhD program supported by the Government of Botswana.

## Data availability

The data supporting the findings of this study are available from BoMRA, the corresponding author, upon reasonable request and within the supplementary information.

## Declarations

### Competing interests

The authors declare no competing interests.

### Ethics approval

The study utilised anonymised data. Following the University of Birmingham’s Research Ethics Committee assessment, no ethical approval was required. The Botswana Ministry of Health, the Health Research Development Committee (HRDC01065), and the Botswana Medicines Regulatory Authority (MRA1/13/1(0001)/25) granted data access.

## Notes

### Competing Interest Statement

The authors have declared no competing interest.

### Author Declarations

The study utilised anonymised data. Following the University of Birmingham Research Ethics Committee assessment no ethical approval was required. The Botswana Ministry of Health, the Health Research Development Committee (HRDC01065) and the Botswana Medicines Regulatory Authority (MRA1/13/1(0001)/25) granted data access.

### Summary of Updates

The ethics approval statement has been updated to The study utilised anonymised data. Following the University of Birmingham Research Ethics Committee assessment no ethical approval was required. The Botswana Ministry of Health the Health Research Development Committee (HRDC01065) and the Botswana Medicines Regulatory Authority (MRA1/13/1(0001)/25) granted data access.

