## Supplementary Information for "Pharmacovigilance of Integrase Strand Transfer Inhibitors in Botswana: Safety Signals, Report Completeness, and Causality Assessment"

Table S1. Two-by-two contingency table.

| | Reports with AE of interest (Y) | All other reports without the AE of interest | Total |
|---|---|---|---|
| Reports for drug of interest (X) | a | b | a + b |
| All other drugs | c | d | c + d |
| Total | a + c | b + d | a + b + c + d |

Table S2. Algorithms used for signal detection.

| Statistic | Formula | Criteria |
|---|---|---|
| BCPNN | $IC = log^2 [(a + 0.5)/ ((a + b)\ (a + c))/ (a + b + c + d)) + 0.5)]$ | $IC_{025} > 0$, $a \geq 3$ |

Notes. BCPNN: Bayesian confidence propagation neural network, IC: information component, $IC_{025}$: lower limit of the 95% confidence interval of the IC. **"a";** number of adverse events of interest attributed to drug X. **"b";** number of other adverse events of drug X, except the adverse event of interest. **"c;"** number of adverse events of interest for all other drugs except for drug X. **"d;"** number of all other adverse events attributed to all other drugs.

Table S3. Disproportionality analysis of cabotegravir and dolutegravir at the preferred term level.

| Active ingredient | Reaction (PT) | *N* | $IC_{025}$ | IC |
|---|---|---|---|---|
| Cabotegravir | Injection site pain | 7 | 1.8 | 3.0 |
| Cabotegravir | Swelling | 4 | 1.0 | 2.8 |
| Cabotegravir | Pain | 8 | 0.7 | 1.9 |
| Dolutegravir | Overdose | 3 | 0.6 | 2.6 |
| Dolutegravir | Rash | 8 | 0.2 | 1.3 |
| Dolutegravir | Medication error | 3 | 0.2 | 2.2 |
| Cabotegravir | Rash | 4 | 0.1 | 1.8 |
| Dolutegravir | Pruritus | 6 | -0.1 | 1.3 |
| Dolutegravir | Neural tube defect | 2 | -0.3 | 2.2 |
| Dolutegravir | Lipodystrophy acquired | 4 | -0.4 | 1.4 |
| Cabotegravir | Pruritus | 3 | -0.4 | 1.7 |
| Dolutegravir | Renal impairment | 8 | -0.4 | 0.7 |
| Dolutegravir | Rash maculo-papular | 2 | -0.5 | 2.1 |
| Cabotegravir | Injection site swelling | 2 | -0.5 | 2.1 |
| Dolutegravir | Renal failure | 11 | -1.2 | -0.2 |
| Cabotegravir | Pain in extremity | 2 | -1.2 | 1.4 |
| Dolutegravir | Weight increased | 2 | -1.4 | 1.2 |
| Dolutegravir | Neuropathy peripheral | 2 | -1.7 | 0.9 |
| Cabotegravir | Dizziness | 3 | -2.1 | 0.0 |
| Raltegravir | Bone development abnormal | 1 | -2.2 | 1.6 |
| Cabotegravir | Abscess limb | 1 | -2.2 | 1.6 |
| Cabotegravir | Fall | 1 | -2.2 | 1.6 |
| Cabotegravir | Thermal burn | 1 | -2.2 | 1.6 |
| Cabotegravir | Administration site discolouration | 1 | -2.2 | 1.6 |
| Cabotegravir | Cheilitis | 1 | -2.2 | 1.6 |
| Dolutegravir | Meningomyelocele | 1 | -2.2 | 1.6 |
| Dolutegravir | Presyncope | 1 | -2.2 | 1.6 |
| Dolutegravir | Myositis | 1 | -2.2 | 1.6 |
| Dolutegravir | Otorrhoea | 1 | -2.2 | 1.6 |
| Dolutegravir | Staphylococcal skin infection | 1 | -2.2 | 1.6 |
| Dolutegravir | Skin depigmentation | 1 | -2.2 | 1.6 |
| Dolutegravir | Lichenification | 1 | -2.2 | 1.6 |
| Dolutegravir | Tongue discolouration | 1 | -2.2 | 1.6 |
| Dolutegravir | Depression | 1 | -2.3 | 1.5 |

| | | | | |
|---|---|---|---|---|
| Dolutegravir | Acne | 1 | -2.3 | 1.5 |
| Dolutegravir | Psychomotor hyperactivity | 1 | -2.3 | 1.5 |
| Dolutegravir | Skin laceration | 1 | -2.3 | 1.5 |
| Dolutegravir | Photodermatosis | 1 | -2.3 | 1.5 |
| Dolutegravir | Vitiligo | 1 | -2.3 | 1.5 |
| Dolutegravir | Rash macular | 1 | -2.3 | 1.5 |
| Dolutegravir | Hallucination | 1 | -2.3 | 1.5 |
| Dolutegravir | Expired product administered | 1 | -2.3 | 1.5 |
| Dolutegravir | Underdose | 1 | -2.3 | 1.5 |
| Dolutegravir | Creatinine renal clearance decreased | 3 | -2.3 | -0.2 |
| Dolutegravir | Product dose omission issue | 1 | -2.3 | 1.5 |
| Cabotegravir | Erectile dysfunction | 1 | -2.4 | 1.4 |
| Dolutegravir | Appetite disorder | 1 | -2.4 | 1.4 |
| Dolutegravir | Libido decreased | 1 | -2.4 | 1.4 |
| Dolutegravir | Blood creatinine | 1 | -2.4 | 1.4 |
| Dolutegravir | Rash papular | 1 | -2.5 | 1.3 |
| Dolutegravir | Skin hyperpigmentation | 1 | -2.5 | 1.3 |
| Dolutegravir | Salivary hypersecretion | 1 | -2.6 | 1.2 |
| Dolutegravir | Pneumonia | 1 | -2.6 | 1.2 |
| Cabotegravir | Lip swelling | 1 | -2.6 | 1.2 |
| Dolutegravir | Stevens-Johnson syndrome | 1 | -2.7 | 1.1 |
| Dolutegravir | Increased appetite | 1 | -2.7 | 1.1 |
| Dolutegravir | Erythema | 1 | -2.8 | 1.0 |
| Dolutegravir | Chronic kidney disease | 1 | -2.8 | 1.0 |
| Cabotegravir | Peripheral swelling | 1 | -3.0 | 0.8 |
| Dolutegravir | Nephropathy toxic | 1 | -3.0 | 0.8 |
| Dolutegravir | Adverse drug reaction | 1 | -3.0 | 0.8 |
| Cabotegravir | Dyspnoea | 1 | -3.1 | 0.7 |
| Dolutegravir | Rash pruritic | 1 | -3.2 | 0.6 |
| Dolutegravir | Muscle spasms | 1 | -3.4 | 0.4 |
| Dolutegravir | Gynaecomastia | 1 | -3.5 | 0.3 |
| Cabotegravir | Fatigue | 1 | -3.6 | 0.2 |
| Cabotegravir | Headache | 2 | -3.7 | -1.1 |
| Dolutegravir | Hepatic enzyme increased | 1 | -3.8 | 0.0 |
| Dolutegravir | Hypoaesthesia | 1 | -4.2 | -0.4 |
| Dolutegravir | Pain in extremity | 1 | -4.3 | -0.5 |
| Dolutegravir | Diarrhoea | 1 | -4.7 | -0.9 |
| Dolutegravir | Cough | 1 | -5.1 | -1.3 |

| | | | | |
|---|---|---|---|---|
| Dolutegravir | Pyrexia | 1 | -5.4 | -1.6 |
| Dolutegravir | Headache | 2 | -5.5 | -2.9 |
| Dolutegravir | Vomiting | 1 | -5.7 | -1.9 |
| Dolutegravir | Pain | 1 | -6.0 | -2.2 |
| Notes: PT; preferred term, IC; information component; $IC_{025}$: lower limit of 95% CI of the IC | | | | |

Table S4. Completeness score by reporter qualification

| Reporter | n | Median | IQR |
|---|---|---|---|
| Other Health Professional | 57 | 0.70 | 0.57-1.00 |
| Unknown | 2 | 0.44 | 0.34-0.54 |
| Pharmacist | 32 | 0.47 | 0.35-0.63 |
| Consumer/Non-Health Professional | 1 | 0.45 | 0.45-0.45 |
| Physician | 20 | 0.56 | 0.42-0.70 |
| IQR: interquartile range | | | |

Table S5. Pairwise Wilcoxon Comparisons with Benjamini-Hochberg (BH) adjustment of completeness scores by reporter

| Comparison | *p*-value (adjusted) |
|---|---|
| Other Health Professionals Pharmacist | p < 0.001 |
| Other Health Professionals vs. Physicians | 0.046 |
| Pharmacist vs Physician | 0.241 |
| Notes: Consumers/non-healthcare professionals were excluded from the analysis | |

Table S6. Linear regression of completeness score versus reporter qualification

| Term | Estimate | Std-Error | T-value | p- value |
|---|---|---|---|---|
| Other Health Professional (reference) | 0.70 | 0.03 | 23.14 | p < 0.001 |
| Pharmacist vs Other Health Professional | -0.21 | 0.05 | -4.08 | p < 0.001 |
| Physician vs Other Health Professional | -0.14 | 0.06 | -2.3 | 0.02 |

Table S7. Linear regression of completeness score versus seriousness

| Seriousness | N | Median | IQR |
|---|---|---|---|
| No | 86 | 0.60 | 0.37 - 0.70 |
| Yes | 25 | 0.72 | 0.57 - 1.00 |
| Unknown | 1 | 0.65 | 0.65 - 0.65 |
| IQR: interquartile range | | | |

Table S8. Logistic regression of completeness score versus seriousness

| Term | OR | 95% CI | z_value | p_value |
|---|---|---|---|---|
| Intercept | 0.06 | 0.013-0.22 | -3.94 | 0 |
| Completeness score | 11.38 | 1.745- 84.97 | 2.47 | 0.013 |
| OR: Odds ratios; CI: confidence intervals | | | | |

Table S9. Agreement statistics between the WHO-UMC system, the Liverpool ADR Causality Assessment Tool (LCAT), and the Naranjo Causality Assessment algorithm.

| Comparison | Kappa | Weighted_Kappa |
|---|---|---|
| LCAT vs WHO-UMC | 0.950696 | 0.985171 |
| LCAT vs Naranjo algorithm | 0.654704 | 0.252971 |
| WHO-UMC vs Naranjo algorithm | 0.627501 | 0.083615 |
| Notes: WHO-UMC: World Health Organization-Uppsala Monitoring Centre system; LCAT: Liverpool ADR Causality Assessment Tool | | |
